# Age and product dependent vaccine effectiveness against SARS-CoV-2 infection and hospitalisation among adults in Norway: a national cohort study, July – November 2021

**DOI:** 10.1101/2022.03.29.22273086

**Authors:** Jostein Starrfelt, Anders Skyrud Danielsen, Eirik Alnes Buanes, Lene Kristine Juvet, Trude Marie Lyngstad, Gunnar Øyvind Isaksson Rø, Lamprini Veneti, Sara Viksmoen Watle, Hinta Meijerink

## Abstract

**Background:** COVID-19 vaccines have been crucial in the pandemic response and understanding changes in vaccines effectiveness is essential to guide vaccine policies. Though the Delta variant is no longer dominant, understanding vaccines effectiveness properties will provide essential knowledge to comprehend the development of the pandemic and estimate potential changes over time.

**Methods:** In this population-based cohort study, we estimated vaccine effectiveness against SARS-CoV-2 infections, hospitalisations, intensive care admissions, and death using Cox proportional hazard models, across different vaccine product regimens and age groups, between 15 July and 31 November 2021 (Delta variant period). Vaccine status is included as a time-varying covariate and all models were adjusted for age, sex, comorbidities, county of residence, country of birth, and living conditions. Data from the entire adult Norwegian population were collated from the National Preparedness Register for COVID-19 (Beredt C19).

**Results:** The overall adjusted vaccine effectiveness against infection decreased from 81.3% (confidence interval (CI): 80.7 to 81.9) in the first two to nine weeks after receiving a second dose to 8.6% (CI:4.0 to 13.1) after more than 33 weeks, compared to 98.6% (CI: 97.5 to 99.2) and 66.6% (CI: 57.9 to 73.6) against hospitalisation respectively. After the third dose (booster), the effectiveness was 75.9% (CI: 73.4 to 78.1) against infection and 95.0% (CI: 92.6 to 96.6) against hospitalisation. Spikevax or a combination of mRNA products provided the highest protection, but the vaccine effectiveness decreased with time since vaccination for all vaccine regimens.

**Conclusions:** Even though the vaccine effectiveness against infection wanes over time, all vaccine regimens remained effective against hospitalisation after the second vaccine dose. For all vaccine regimens, a booster facilitated recovery of effectiveness. The results from this support the use of heterologous schedules, increasing flexibility in vaccination policy.

**Funding:** no external funding

## Background

Since the start of the COVID-19 pandemic, various COVID-19 vaccines have been approved for Emergency Use Listing/Authorization (EUL/EUA), including Comirnaty (Pfizer/BioNTech; BNT162b2), Spikevax (Moderna; mRNA-1273), Vaxzevria (AstraZeneca; ChAdOx nCoV-19; AZD1222), and Janssen (Johnson & Johnson; Ad26.COV2.S). Both vaccine efficacy estimates from randomised controlled trials and vaccine effectiveness estimates from observational studies in the first months after the vaccine roll-out showed strong protection against both infection and severe disease.^1-6^ However, effectiveness may differ between product types and against different virus variants, as well as be affected by dose intervals or population structure (age distribution, risk groups).^7-13^ Many countries have adopted flexible policies allowing “mixing and matching” of vaccines (heterologous regimens) during SARS-CoV-2 vaccination campaigns, in response to supply constraints, policy changes or rare but severe side effects associated with the vector-based vaccines.^14,15^ Combining the vector-based vaccines, such as Vaxzevria, with an mRNA vaccine increases the vaccine effectiveness to a level comparable with mRNA regimens.^7,16-18^ Nonetheless, a possible waning of vaccine-induced immunity could result in lower vaccine effectiveness over time.^19,20^ Recently published data suggest reduced protection against SARS-CoV-2 infection in all age groups six months after the completion of a primary vaccination regimen, and also, a small decrease against severe disease in certain groups.^21^

On 27 December 2020, Norway started COVID-19 vaccination, initially targeted towards elderly (>65 years) and risk groups. Of those ≥18 years, 88% had received at least two vaccine doses by 5 December 2021. Vaxzevria was included in the Norwegian national vaccine programme until 11 March 2021; those who received one dose were offered a second dose with an mRNA vaccine. Since September 2021, a booster dose has been recommended, initially prioritising those above 65 years and risk groups, including health care workers. From early February 2021, the Alpha variant (B1.1.7) was the dominant circulating strain in Norway, being replaced by the Delta variant (B.1.617.2) by July and Omicron by December 2021.^5,7,8^

Understanding the changes in vaccines effectiveness over time, the impact of giving boosters, and differences between vaccine types, is essential to guide vaccine implementation and policies. Even though the Delta variant is no longer dominant in many regions, understanding these properties will provide essential knowledge that can be used to understand the development of the pandemic and estimate potential changes over time. The purpose of this study is to quantify and compare the vaccine effectiveness against infection, disease, and death achieved in the Norwegian population during the Delta epidemic considering time since vaccination, vaccine type and age groups.

## Methods

### Study population

For this population-based cohort study, we collated data from the Norwegian National Preparedness Register for COVID-19 (Beredt C19) (Supplementary table S1), which contains individual-level data from national central health registries, national clinical registries, and other national administrative registries. We included all adults (≥18 years by the end of 2021) with a valid national identity number and registered in the National Population Registry (NPR) as living in Norway. To remove non-standard vaccination histories, we removed individuals with more than three doses before the end of the study period (1 348 individual) and excluded individuals for which the interval between first and second dose was shorter than the recommended minimum intervals (678 individuals), and censored those with a third dose registered before the recommended 120 days of the second dose (17 497 individuals). The recommended minimum interval between first and second dose was based on the vaccine type given as the first dose; 19 days for Comirnaty, 22 days for Spikevax, and 21 days for Vaxzevria. We only included individuals who had received either of the three vaccines that are part of the Norwegian vaccination programme (Comirnaty, Spikevax or Vaxzevria), excluding a further 6 257 individuals. Finally, 67 individuals admitted to hospital with COVID-19 without a corresponding match in the database for the time of positive test were excluded. Data were extracted from the registries on 15 February 2022.

### Definitions

SARS-CoV-2 infection is defined as a positive SARS-CoV-2 PCR test reported to the Norwegian Surveillance System for Communicable Diseases (MSIS) registry. Individuals who were hospitalised or admitted to an ICU with COVID-19 are registered in the Norwegian Intensive Care and Pandemic Registry (NIPaR). We included all hospitalisation where COVID-19 was registered as the primary diagnosis for admission and ICU admission of individuals who tested positive for SARS-CoV-2 and were admitted to an ICU (length of stay ≥24 hours), required mechanical ventilatory support (invasive or non-invasive), or persistent administration of vasoactive medication. All COVID-19 associated deaths are defined as anyone with a positive SARS-CoV-2 PCR test who died with COVID-19 reported on the death certificate in the Cause of Death Register (DÅR), or those notified directly to MSIS. We use testing date as time of infection (positive PCR test) and vaccination status is determined at time of infection for all outcomes. Individual vaccination histories were generated from the Norwegian Immunisation Registry (SYSVAK), and categorised into the following vaccination statuses:

- Unvaccinated: unvaccinated up to seven days before the first dose
- 1^st^ dose: ≥21 days after first vaccine dose up to seven days after second vaccine dose
- 2^nd^ dose: >7 days after the 2^nd^ dose, divided in period of eight weeks
- 3^rd^ dose (booster): >7 days after a vaccine dose given 120 days or more after completion of the primary vaccine regimen

The vaccine regimens included were Comirnaty, Spikevax, heterologous mRNA regimen, Vaxzevria, or Vaxzevria in combination with an mRNA vaccine, all with or without an mRNA booster. The period between seven days before the first vaccine dose until 21 days after the first dose was included as a separate status not reported, since vaccination was postponed if individuals showed signs of infection which could potentially bias infection rates for both unvaccinated and partially vaccinated if included in the adjacent vaccination statuses. Similarly, individuals were also included as a separate status and not reported for the first seven days after receiving the third dose. Individuals with a SARS-CoV-2 infection registered prior to 1 June 2021 were included as a separate category (see below).

To adjust for confounding, several covariates were included in our analysis. When stratifying by age, we used the categories 18 to 44 years, 45 to 64 years, and 65 years or older. For adjustment in other models, we used 10-year age bands (NPR). County of residence (NPR) was included as both infection rates and speed of vaccine rollout has varied across Norway. We included country of birth (NPR; Norway, abroad or unknown) and crowded living conditions (Statistics Norway; crowded, not crowded, or unknown) since both are associated with vaccine coverage and risk of infection. Individuals with pre-existing medical conditions associated increased risk of severe COVID-19 illness were prioritised for vaccination and this covariate was also included in the adjusted model. Missing values were considered as a separate category for each of the variables where relevant. More details on the data sources, and variables can be found in the supplementary information as well as the number of infections, hospitalisations, and vaccination status over time (figures S1 and S2).

### Data analyses

We estimated the vaccine effectiveness using Cox proportional hazards models on an open cohort, using vaccine status as time-varying covariate for all individuals included in the statistical software *R*^*18*^. We included all SARS-CoV-2 infections reported from 15 July until 30 November 2021, the period in which the delta variant was dominating in Norway.^10^ We right-censored individuals at the time of an event (SARS-CoV-2 infection, hospitalisation, ICU admission or death associated with COVID-19), time of death (all cause) or end of follow-up period (30 November). During the study period, the registries in Norway only report re-infections if six months or more since last positive test, individuals registered with an infection prior to 15 July 2021 entered the dataset 180 days since positive test with status ‘previously infected’.

Vaccine effectiveness is defined as 100*(1–β), where β represents the hazard ratio associated with a particular vaccine status. For crude vaccine effectiveness estimates, we only used vaccine status as a time-varying covariate (supplementary analyses). For adjusted estimates, we implemented stratified analyses using *strata(variable)* in the *survival-*package^17^, i.e. that the impact of the adjustment variables can be non-proportional.

To estimate specific vaccine effectiveness for age groups and vaccine product regimens, we use independent Cox-models while still adjusting for the remaining covariates as *strata*. Vaccine status was factored either by combining all vaccine types (thus assuming similar effectiveness across vaccines) to estimate a population level vaccine effectiveness of the vaccination program in Norway or by implementing vaccine status as the combination of vaccine type and vaccine status. Due to the smaller numbers and partial exclusion of Vaxzevria from the vaccination program in Norway, all individuals who received a dose of Vaxzevria were censored at time of the dose in the product specific analyses. Models were also run excluding all unvaccinated individuals who have never had a recorded SARS-CoV-2 PCR test in Norway, results from these models can be found in the supplementary materials (subcohort 1).

## Results

In total, 75 303 were diagnosed with SARS-CoV-2 infection, 1 438 were hospitalised with COVID-19 as main cause for admission, 289 were admitted to the ICU, and 331 died with COVID-19 between 15 July and 30 November 2021. Characteristics of the study population and by outcome can be found in Table 1. Overall vaccine effectiveness against infection was estimated at 24 .7 % (confidence interval/CI: 22 .7 to 26 .7) after the first dose, 65 .2 % (CI: 64 .6 to 65 .9) after the second dose and 84.8 % (CI: 83.3 to 86.3) after the third dose. Previous infections reduced the probability of infection by 93.5 % (CI: 92.7 to 94.2).

**Table 1:** Study population

Characteristics of study population and by outcomes of interests; SARS-CoV-2 infections, hospitalisation, ICU admission and death, Norway, 15 July – 30
|  |  | Total study population |  | SARS-CoV-2 infection |  | Hospitalisation |  | ICU admission |  | COVID-19 deaths |  |
| --- | --- | --- | --- | --- | --- | --- | --- | --- | --- | --- | --- |
|  |  | n | % | n | % | n | % | n | % | n | % |
| All | - | 4 301 995 | - | 74 371 | - | 1 429 | - | 290 | - | 331 | - |
| Age groups | 18-24 years | 457 238 | 10.6 | 11 499 | 15.5 | 24 | 1.7 | 1 | 0.3 | 0 | 0.0 |
|  | 25-34 years | 747 249 | 17.4 | 16 013 | 21.5 | 113 | 7.9 | 16 | 5.5 | 2 | 0.6 |
|  | 35-44 years | 705 460 | 16.4 | 17 131 | 23.0 | 190 | 13.3 | 40 | 13.8 | 2 | 0.6 |
|  | 45-54 years | 735 420 | 17.1 | 14 050 | 18.9 | 206 | 14.4 | 46 | 15.9 | 6 | 1.8 |
|  | 55-64 years | 653 259 | 15.2 | 7 440 | 10.0 | 202 | 14.1 | 63 | 21.7 | 22 | 6.6 |
|  | 65-74 years | 540 898 | 12.6 | 4 604 | 6.2 | 214 | 15.0 | 52 | 17.9 | 38 | 11.5 |
|  | 75-84 years | 335 628 | 7.8 | 2 557 | 3.4 | 301 | 21.1 | 59 | 20.3 | 103 | 31.1 |
|  | ≥85 years | 126 843 | 2.9 | 1 077 | 1.4 | 179 | 12.5 | 13 | 4.5 | 158 | 47.7 |
| **Sex** | Male | 2 160 307 | 50.2 | 37 098 | 49.9 | 791 | 55.4 | 196 | 67.6 | 185 | 55.9 |
|  | Female | 2 141 688 | 49.8 | 37 273 | 50.1 | 638 | 44.6 | 94 | 32.4 | 146 | 44.1 |
| **Underlying conditions** | No risk group | 3 401 381 | 79.1 | 62 802 | 84.4 | 711 | 49.8 | 129 | 44.5 | 78 | 23.6 |
|  | Medium risk | 788 954 | 18.3 | 10 247 | 13.8 | 524 | 36.7 | 115 | 39.7 | 180 | 54.4 |
|  | High Risk | 111 660 | 2.6 | 1 322 | 1.8 | 194 | 13.6 | 46 | 15.9 | 73 | 22.1 |
| **Country of birth** | Norway | 3 202 876 | 74.5 | 46 501 | 62.5 | 791 | 55.4 | 170 | 58.6 | 247 | 74.6 |
|  | Outside Norway | 802 615 | 18.7 | 25 579 | 34.4 | 506 | 35.4 | 103 | 35.5 | 34 | 10.3 |
|  | Unknown | 296 504 | 6.9 | 2 291 | 3.1 | 132 | 9.2 | 17 | 5.9 | 50 | 15.1 |
| **Crowding** | Yes | 336 777 | 7.8 | 11 953 | 16.1 | 175 | 12.2 | 29 | 10.0 | 7 | 2.1 |
|  | No | 3 728 263 | 86.7 | 57 206 | 76.9 | 1 154 | 80.8 | 238 | 82.1 | 299 | 90.3 |
|  | Unknown | 236 955 | 5.5 | 5 212 | 7.0 | 100 | 7.0 | 23 | 7.9 | 25 | 7.6 |

November 2021

### Vaccine effectiveness since time of vaccination

The adjusted vaccine effectiveness against infection was high in the first period (two to nine weeks) after the second dose (81.3 %, CI: 80.7 to 81.9). Effectiveness waned with time since vaccination and more than 33 weeks after receiving the second dose the effectiveness against infection was 8.6 % (CI: 4.0 to 13.4). Similarly, the effectiveness against hospitalisation was 98.6 % (CI: 97.5 to 99.2) in the period right after receiving the second dose, decreasing to 66.6 % (CI: 57.9 to 73.6) after more than 33 weeks. For admission to intensive care unit and death, not enough events in the first period following the second dose had occurred to reliably estimate a vaccine effectiveness, but in the following period (10 to 17 weeks), vaccine effectiveness was 96.9 % (CI: 94.7 to 98.1) and 93.4 % (CI: 85.4 to 97.0) against ICU and death respectively, with a less pronounced reduction over time for ICU admissions (86.7%, CI: 73.9 to 93.2 after more than 33 weeks) than for death (68.6%, CI: 55.4 to 77.9). One dose provided little protection against infection (30.0 %, CI: 28.2 to 31.9), but did protect against hospitalisation (79.4 %, CI: 73.0 to 84.2), ICU admission (92.4 %, CI: 80.9 to 97.0) though not death (46.9 %, CI: -0.2 to 71.9). The vaccine effectiveness against infection after receiving a third dose (75.9 %, CI: 73.4 to 78.1) was similar to the effectiveness in the initial two to nine weeks after the second dose (81.3 %, CI: 80.7 to 81.9). For those with a previous reported infection (>6 months prior), the protection against infection was 93.1 % (CI: 92.3 to 93.9), whereas too few events among those with a reported prior infection were reported to estimate effectiveness against hospitalisation. Vaccine effectiveness for all outcomes split by time are shown in Figure 1 (details S2-S5).

**Figure 1.**
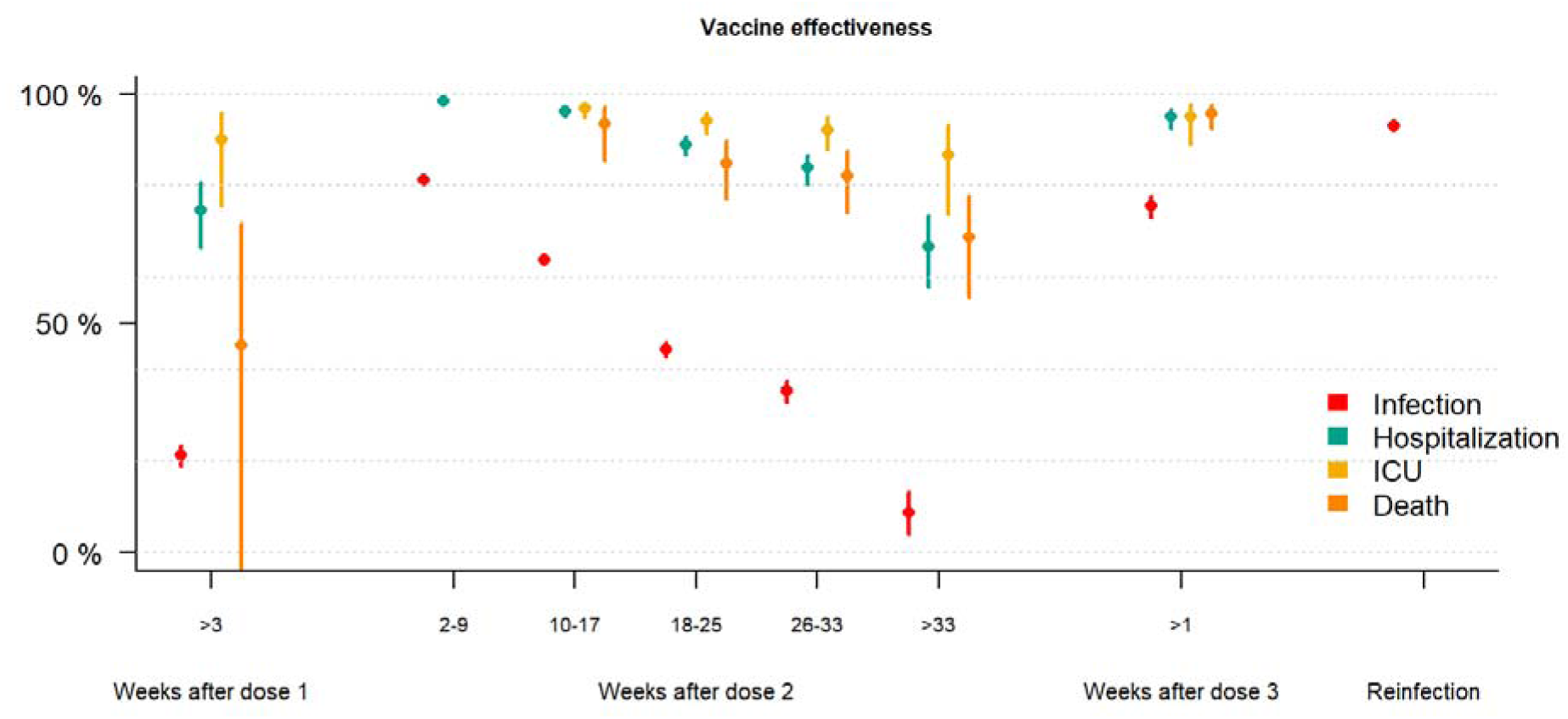
Adjusted vaccine effectiveness against infection (red), hospitalisation (blue), ICU admission (yellow) and COVID-19 associated deaths (orange) for Norwegian adults using data from 15 July - 30 November 2021. *Adjusted for age, sex, comorbidities, county of residence, country of birth, and living conditions*.

### Vaccine effectiveness by age

Vaccine effectiveness against infection was highest in two to nine weeks after the second dose among 18-to 44-year-olds (83.2 %; CI: 82.6 to 83.8) compared to 45-to 64-year-olds (75.6 %; CI: 74.1 to 77.0) and those over 64 years (74.9 %; CI: 67.2 to 80.7). No significant protection was found more than 33 weeks after the second dose: the estimated effectiveness against infection was 5.2 % (CI: -1.9 to 11.8) for 18-to 44-year-olds, 0.5 % (CI: -9.4 to 9.5) among 45-to 64-year-olds and 8.4 % (−2.8 to 18.5) among those over 65 years (Figure 2, Table S6). Among those with a reported prior infection, protection against infection was 92.6 % (CI: 91.5 to 93.5), 95.1 % (CI: 93.6 to 96.3) and 89.0 % (CI: 81.3 to 93.5) for 18- to 45- year-olds, 45- to 64-year-olds and over 65-yea-olds respectively (Figure 2). The protection against hospitalisation was already significant after one dose in all age groups: 82.9 % (CI: 73.0 to 89.2) among 18- to 44-year-olds, 71.4 % (56.2 to 81.4) among 45- to 64-year-olds and 56.5 % (CI: 24.6 to 74.9) among those over 65 years. The effectiveness against hospitalisation decreased less with time than protection against infection. For 45- to 64-year-olds the effectiveness was 99.1 % (CI: 97.7 to 99.6) in two to nine weeks after the second dose, compared to 65.3 % (CI: 33.3 to 82.0) more than 33 weeks after the second dose. Similarly, among those above 65 years, the protection waned from 93.6 % (CI: 89.9 to 96.0) to 61.9 % (CI: 50.1 to 70.9) (Figure 2, supplementary tables S6 and S7). Receiving a third dose increased vaccine effectiveness against hospitalisation to 85.4 % (CI: 65.3 to 93.9) amongst 45- to 64-year-olds and 95.3 % (CI: 92.6 to 97.0) for over 65-year-olds; the number of events were too small for 18- to 44-year-olds (Figure 2, supplementary tables S6 and S7).

**Figure 2.**
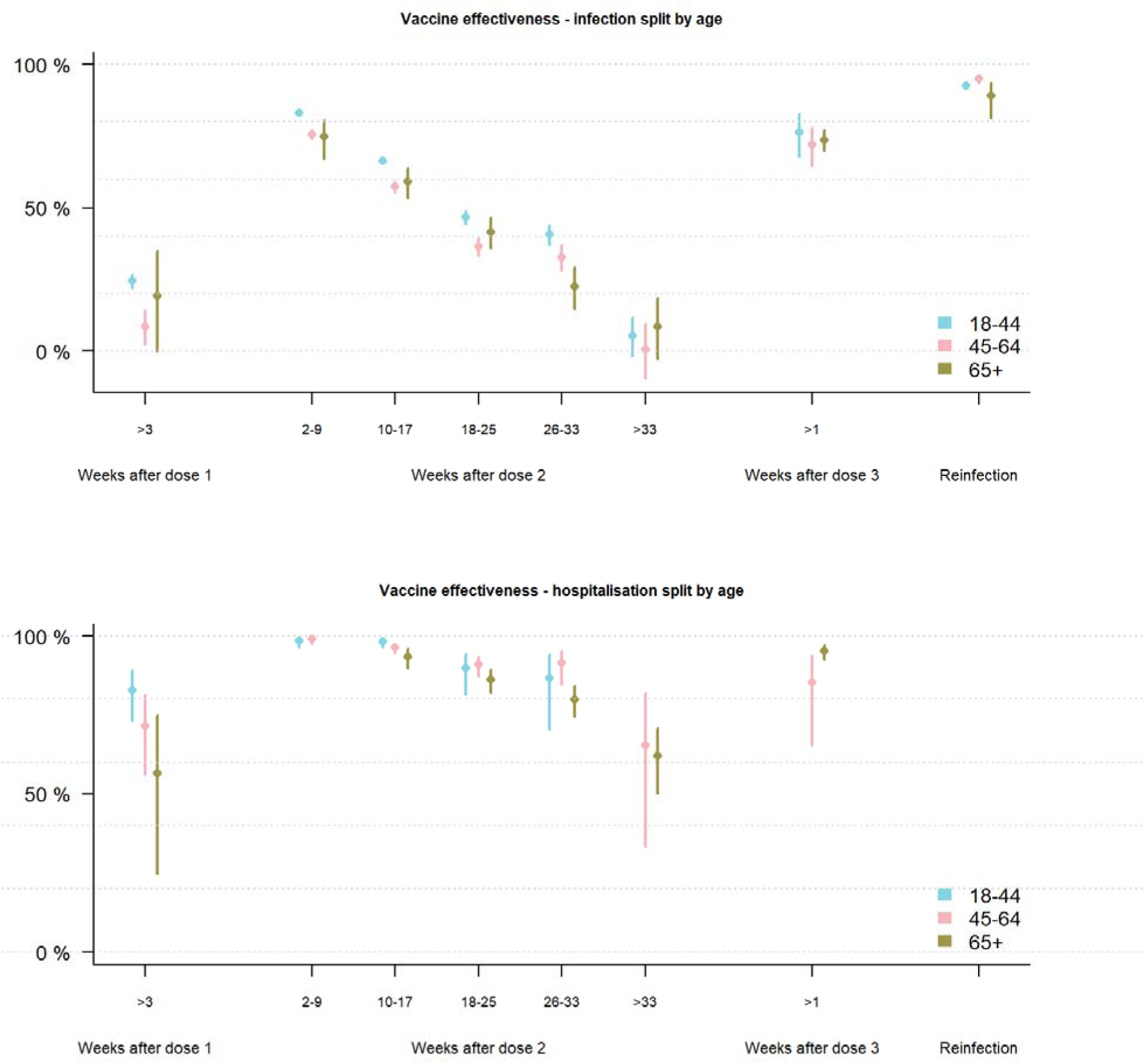
Adjusted vaccine effectiveness against infection (A) and hospitalisation (B) for age 18-44 years (blue), 45 to 64 (pink) and 65+ years (green) among Norwegian adults using data from 15 July – 30 November 2021. *Adjusted for age, sex, comorbidities, county of residence, country of birth, and living conditions*.

### Vaccine effectiveness by product regimen

When stratifying by product regimen, individuals who received two doses of Spikevax (86.6%; CI: 85.6 to 87.6) or a heterologous mRNA regimen (84.1 %; CI: 83.2 to 85.0) had a higher estimated vaccine effectiveness against infection than those who received two doses Comirnaty (77.7 %; CI: 76.8 to 78.5) in two to nine weeks after the second dose (Figure 3, table S8). All product regimens showed waning of vaccine effectiveness against infection. The vaccine effectiveness against infection after receiving the third dose was highest for those who received two doses of Spikevax followed by a booster with Comirnaty (87.1 %; CI: 80.1 to 91.6) or Spikevax (84.9 %; CI: 71.8 to 91.9), compared to 75.3% (CI: 72.5 to 77.8) and 68.2 % (CI: 57.6 to 76.1) for those who received two doses of Comirnaty followed by a booster with Comirnaty or Spikevax respectively. The vaccine effectiveness against hospitalisation was high after one dose (77.1% and 75.3%) for both Spikevax and Comirnaty, as well as one to 32 weeks after the second dose (range 81.8 to 97.5%). There were only five hospital admissions among those who received heterologous mRNA vaccination during our study period and therefore vaccine effectiveness against hospitalisation since time of vaccination could not be estimated. Among those who received a booster dose, vaccine effectiveness against hospitalisation could only be estimated for those with primary regimen of Comirnaty and protection was high 95.6 % (CI: 93.1 to 97.2) for those receiving a Comirnaty booster, but slightly lower but more uncertain for those receiving Spikevax (73.5%; CI: 45.7 to 87.1) (Figure 3, table S9).

**Figure 3.**
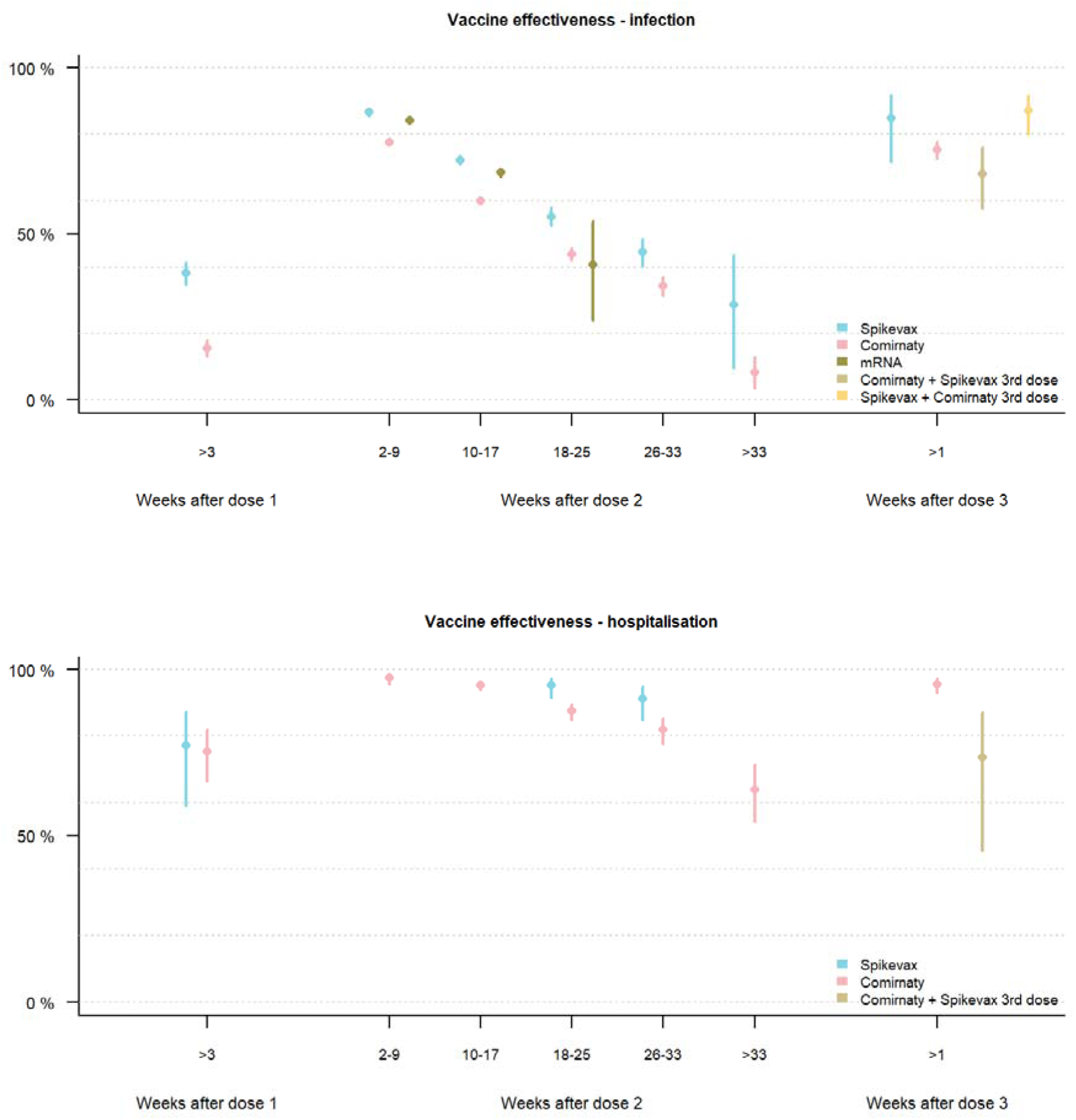
Adjusted vaccine effectiveness against infection (A) and hospitalisation (B) per vaccine product regimen (Spikevax (blue), Comirnaty (pink) or mixed mRNA primary regimen (dark green) and mixed booster (light green and yellow) among Norwegian adults using data from 15 July – 30 November 2021. *Adjusted for age, sex, comorbidities, county of residence, country of birth and living conditions. mRNA includes a combination of one dose Spikevax and one dose Comirnaty; only few individuals who received a heterologous primary regimen were eligible for a booster during the study period and are therefore not included*

## Discussion

Our analyses showed strong protection against SARS-CoV-2 infection in the first period after two or more vaccine doses for both homologous and heterologous mRNA regimens. However, for all product regimens and age groups, the vaccine effectiveness against infection waned over time. Vaccine effectiveness against hospitalisation was high for all product regimens and age groups, with limited waning with time since vaccination.

Our findings are consistent with other studies investigating waning of vaccine effectiveness, as summarised through a systematic review by Feikin et al.^21^ Most observational studies employ a test-negative study design, while some are register-based cohort studies like ours.^19,25-33^ We reported large waning of effectiveness against SARS-CoV-2 infection, which could have several explanations. First, we have a longer follow-up time than previously published studies. Second, we use data from a high-quality population register covering an entire national population in a country with widespread and free testing irrespective of symptoms. After vaccination, individuals might modify their behaviour based on evaluating risk and change propensity for testing, or unvaccinated could become indirectly protected by an increasing population immunity. In addition, unrecorded prior infections among unvaccinated may reduce the detected risk among unvaccinated. The estimates from this study are possibly conservative; excluding those reported as unvaccinated who have never been tested resulted in higher vaccine effectiveness for all outcomes (supplemental tables S2-S5, subcohort 1). Our findings are in line with immunological findings suggesting that antibody titres wane over time.^20,34^ Even though antibody-mediated immunity may wane and require time to reactivate upon infection, the fact that vaccine effectiveness against severe disease remains high is consistent with the induction of cell-mediated immunity.^35^ While groups at risk for more severe outcome form a disproportionate part of those hospitalized, admitted to ICU or death (Table 1), an analysis of a subcohort excluding all risk groups showed similar estimates of vaccine effectiveness (supplemental tables S2-S5, subcohort 2), which indicates that the vaccines reduce the probabilities of infection and hospitalisation similarly across risk-groups.

To our knowledge, this is among the few studies to report on the vaccine effectiveness of heterologous mRNA vaccine regimens. As heterologous vaccine regimens were accepted in Norway from June 2021, recipients of a combination of mRNA vaccines are predominantly younger and healthier. However, these results are also maintained across age groups and are in line with a test-negative case control study from Canada.^28^ In addition, we show that Spikevax shows a slightly higher vaccine effectiveness against infection than Comirnaty, as has also been reported by others.^9,37,38^

During our study period, many aspects of the disease dynamics changed, and changes also occurred simultaneously. This leads to difficulties in correctly identifying the mechanisms and factors driving the system. Even though we attempt to control for these factors in the analyses, residual confounding cannot be excluded. Furthermore, testing intensity changed over time, especially with the introduction of self-administered rapid antigen tests after last summer. Even though testing capacity is high in Norway, all individuals infected with SARS-CoV-2 have not been detected, which could affect the estimates since the proportion of unidentified cases may differ dependent on age as well as on vaccine status.^29^ Additionally, the estimated vaccine effectiveness can be affected by number and types of contacts.^30^ As previously mentioned, the vaccine effectiveness could be underestimated if getting vaccinated results in behavioural changes associated with higher risk of exposure. We are not able to model these effects in our analyses. Nevertheless, the estimated population-level effects can provide a reasonable estimate of the individual-level vaccine efficacy since the total attack rate during the study period was small.^30^

The ability to link data collected via national registries is a great advantage and allows us to estimate population-wide vaccine effectiveness. However, some limitations of register-based data should be considered when interpreting the results. Data in these registries are not collected for the purpose of this study, and therefore the focus on level of detail, error checking and precision in the available data is not guided by the current study, as would be the case for independent data gathering. For instance, while vaccines administered as part of the Norwegian vaccination program should be in our dataset, it is not unlikely that some have received vaccines outside Norway and not reported them to the Norwegian register (SYSVAK). While limitations in register-based data are important caveats – our cohort study encompassing the whole Norwegian adult population indicates that vaccine effectiveness against severe disease is high among vaccinated individuals. Our estimates remain qualitatively the same for protection against infection and severe disease when splitting by age groups, indicating that the confounding effect of factors that are relatively constant within age-groups introduce little bias in our adjusted models.

Appropriate prioritisation and planning of vaccine campaigns is integral for combating COVID-19 and is only possible with updated knowledge on vaccine effectiveness of realistic vaccination regimens achieved in large populations. For our study the overall protection (i.e. a weighted mean of vaccine effectiveness over time) increase through the initial period with a peak of right below 60% on the 21st of September (Supplemental figure S3). Coupling VE-estimates with cohort fractions over time can yield valuable information on the general level of protection in a population and timing and prioritisation of vaccine roll-out. Our study among adults in Norway indicate a high vaccine effectiveness against both infection and hospitalisation with both homologous and heterologous mRNA regimens. Even though the effectiveness against infection declines with time since vaccination, the protection against severe disease remained high. The results support the use of heterologous regimens, increasing flexibility in vaccination policy.

## Conclusions

During the Delta-phase of the COVID-19 epidemic in Norway, vaccine effectiveness against infection clearly waned over time, however, all vaccine regimens remained effective against hospitalisation after the second vaccine dose. For all vaccine regimens, a booster facilitated recovery of effectiveness. The results from this support the use of heterologous schedules, increasing flexibility in vaccination policy.

## Supporting information

STROBE Statement

## Data Availability

Legal restrictions prevent the researchers from sharing the dataset used in the study.

## Declarations

### Ethics approval

Ethical approval was granted by Regional Committees for Medical and Health Research Ethics (REC) Southeast (reference number 122745). The Norwegian Institute of Public Health has performed a Data Protection Impact Assessment (DPIA) for Beredt C19.

### Consent for publication

Not applicable.

### Availability of data and materials

The datasets analysed during the current study come from the national emergency preparedness registry for COVID-19 (Beredt C19), housed at the Norwegian Institute of Public Health. The preparedness registry comprises data from a variety of central health registries, national clinical registries, and other national administrative registries. Legal restrictions prevent the researchers from sharing the dataset used in the study. However, external researchers are freely able to request access to linked data from the same registries from outside the structure of Beredt C19, as per normal procedure for conducting health research on registry data in Norway (https://www.helsedata.no). Further information on the preparedness registry, including access to data from each data source, is available at https://www.fhi.no/en/id/infectiousdiseases/coronavirus/emergency-preparedness-register-for-covid-19/ Code and model results in summary form from R is available from the author.

### Authors’ contribution

JS and HM developed the concept and design for the study. JS performed data analyses, TL, AD, and GR verified the underlying data and model code. JS, AD and HM interpreted the data and drafted the manuscript with inputs from SW, GR, TL, EB, LV and LJ. All authors had the opportunity to request data access. All authors contributed to and approved the submitted version.

## Acknowledgements

The authors greatly appreciate the efforts of all health professionals in vaccinating over 4 million Norwegians and performing over eight million PCR tests, and in addition managing to report these events to form the data for this analysis. We also acknowledge the efforts of employees at hospitals around Norway in the reporting of timely and complete data to the Norwegian Intensive Care and Pandemic Registry, as well as staff in the registry administration. Additional appreciation is given to all contributors to the development and inner workings of Beredt C19.

