## Supplementary material for "Age and product dependent vaccine effectiveness against SARS-CoV-2 infection and hospitalisation among adults in Norway: a national cohort study, July – November 2021"

### Data sources

In this study, we use data from several national registries that were individually linked in the Norwegian preparedness registry (Beredt C19). The data sources and variables used are shown in Table S1

Table S1**. Data sources in the Norwegian preparedness registry (Beredt C19) used in this study and variables retrieved from each source**

| **Norwegian Abbreviation** | **Full name of data source** | **Information obtained** |
| --- | --- | --- |
| DSF | The National Population Register | Age, Sex, County of residence*, Country of birth, Date of death |
| SYSVAK | The National Immunisation Register | Date of vaccination, Vaccine product type |
| NIPaR | Norwegian Intensive Care and Pandemic Registry | Date of hospitalisation, COVID-19 as main cause of admission, Date of ICU admission |
| MSIS | The Surveillance System for Infectious Diseases | Date of sample of SARS-CoV-2 positive test, Date of COVID-19 associated death |
| SSB | Statistics Norway | Crowding (see definition below) |
| Beredt C19 risikogrupper | Table prepared in Beredt C19  Source: Norwegian Patient Registry (NPR): individual level data from all public specialist health-care services in Norway. | Defines risk groups (see definition below) |

* The county of residence was updated in January 2022, which might lead to some errors for individuals who have moved the last half year

Crowding^41^: Individuals are considered to live in crowded conditions if the number of rooms is lower than the number of residents or one resident lives in one room, and the number of square metres (P-area) is below 25 sq. m. per person. If the number of rooms or the P-area is not specified, a household will be regarded as crowded if one of these criteria is met.

Risk groups: Some underlying medical conditions increase the risk of severe COVID-19 outcomes, regardless of age. These individuals have been prioritised in the vaccination programme in Norway. The underlying comorbidities that have been defined as increasing the risk of severe COVID-19 are divided into two groups:

High risk: people with diseases/conditions that carry a high risk of severe COVID-19:

- Organ transplant
- Immunodeficiency
- Haematological cancer in the last five years
- Other active cancers
- Neurological or neuromuscular diseases that cause impaired cough or lung function (e.g., ALS and cerebral palsy)
- Chronic kidney disease, or significant renal impairment.

Medium risk: people with diseases/conditions that entail a moderate risk of severe COVID-19:

- Chronic liver disease or significant hepatic impairment
- Diseases requiring immunosuppressive therapy
- Diabetes
- Chronic lung disease including cystic fibrosis and severe asthma which have required the use of high dose inhaled or oral steroids within the past year
- Obesity with a body mass index (BMI) of ≥35 kg/m2
- Dementia
- Chronic heart and vascular disease (with the exception of high blood pressure) and stroke

### The COVID-19 epidemic in Norway July-Nov 2021

Figure S1 shows the number of infections and hospitalisation and Figure S2 shows the changing proportion of the different vaccine status categories from July 15^th^ to November 30^th^, 2021.

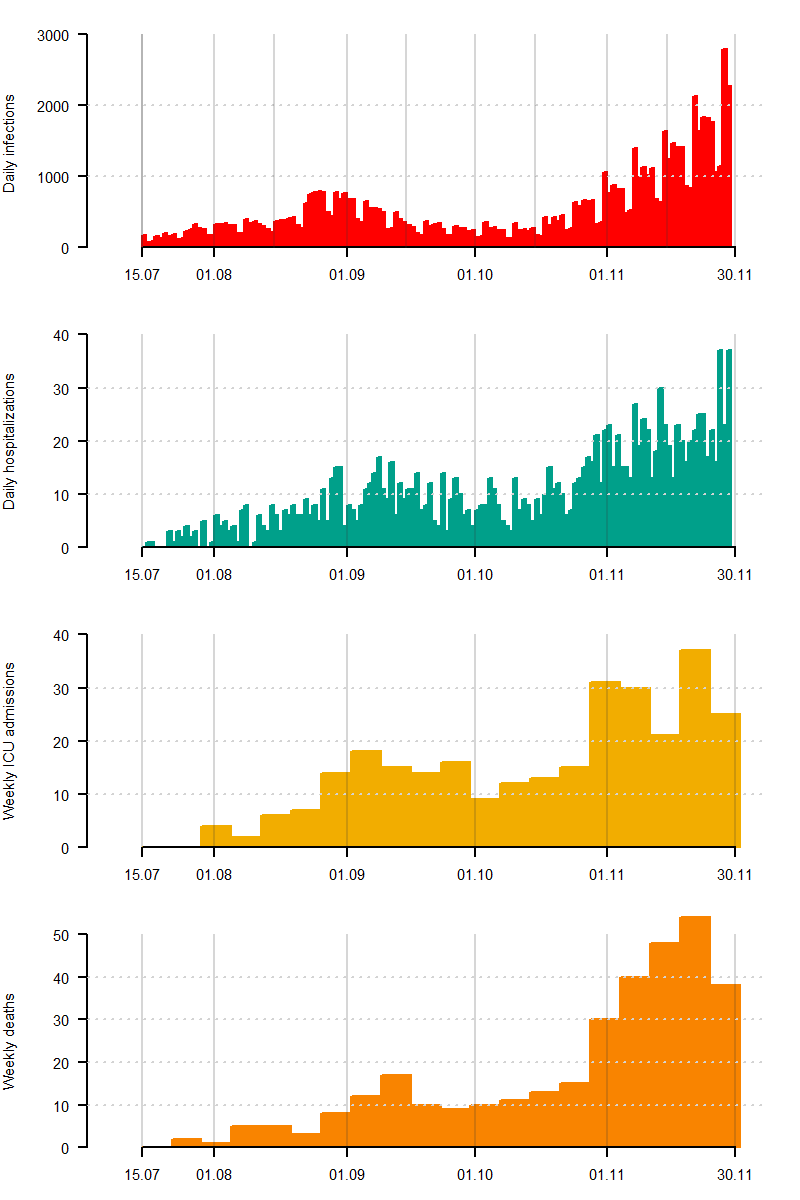

Figure S1. **Daily reported cases of SARS-CoV-2 infections (top, red) and number of hospitalisations (upper middle panel, green), and weekly admissions to intensive care unit (ICU, lowed middle, yellow) and deaths (lower, orange) with Covid listed in death certificate in Norway, 15^th^ of July 2021 - 30^th^ of November 2021**

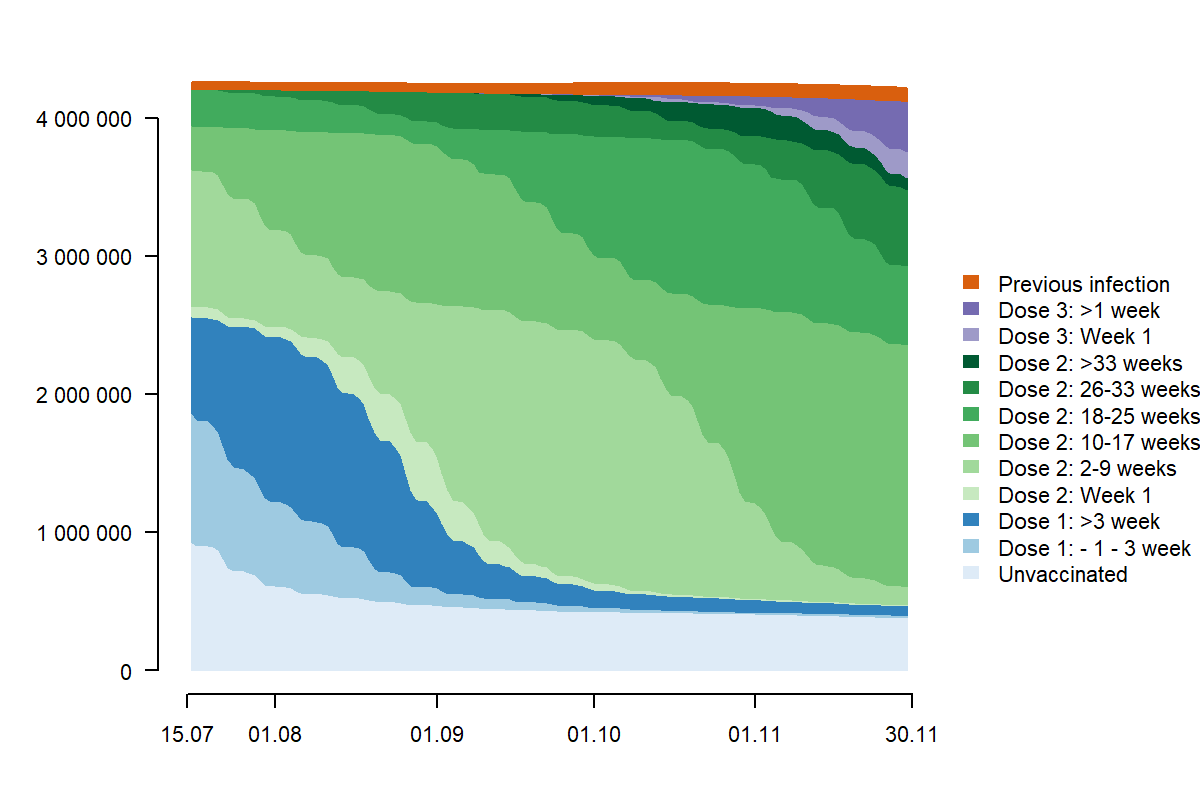

Figure S2. **Distribution of vaccine statuses over time in the period 15th of July – 30th of November.** The initial cohort had 4 253 872 individuals, with a maximum of 4 253 982 on 19th of July, ending with 4 214 429 on 30th of November.

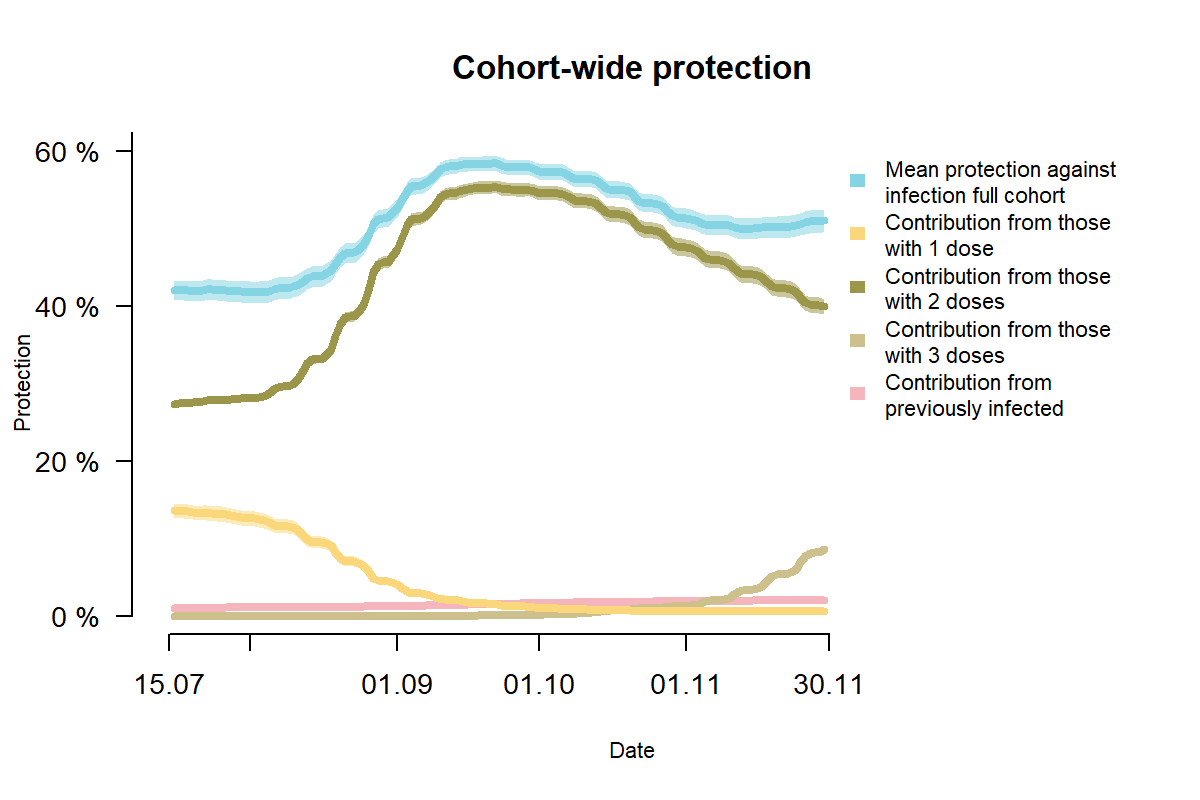

Figure S3**. Cohort-wide level of protection against infection over time. On any individual day the fraction of individuals belonging to the different vaccination categories (see Figure S2) were multiplied with the estimate vaccine effectiveness in the main model without splitting by age and product-types (Table S2).** In essence the figure shows the weighted mean vaccine effectiveness over time (blue), with contributions to the overall mean from those with 1 dose (yellow, including period right before and 1 week after 1st dose), individuals with 2 doses (green, including 1st week after 2nd dose), those with 3rd dose (light green, including 1st week after 3rd dose) and those included as previously infected (pink). Note that these contributions are also weighted, i.e. the reason why the pink line for the previously infected is low is that they contribute very little to the overall protection because they constitute a very little part of the cohort, though at the group level they are estimated to have very high protection.

### Modelling vaccine effectiveness

Using Cox proportional hazard models, we estimated the COVID-19 vaccine effectiveness against SARS-CoV-2 infection, hospitalisation, ICU admission, and death associated with COVID-19. We ran three sets of model structures:

- Unadjusted models only using vaccine status as a time-varying covariate. This yields crude vaccine effectiveness estimates.
- Adjusted models with vaccine status as well as age, sex, crowding, country of birth, county of residence and risk group as fixed covariates assuming proportional impact on hazard rates. These were performed to test for impact of covariates.
- Adjusted models with all covariates, except vaccine status, implemented as stratification variables. This allows each combination of covariate groups to have their own baseline hazard rate, while still assuming that vaccination leads to a proportional change in risk of infection, i.e. a less restrictive assumption on the impact of confounders.

For adjusted models where covariates were included as factors whose effect was assumed proportional and directly estimated, for all covariates at least some levels had significant impacts when in models for all adults and not split by vaccine type. We therefore report adjusted vaccine effectiveness estimates in the manuscript from the third model structure allowing impacts of all covariates except vaccine status to possibly be non-proportional.

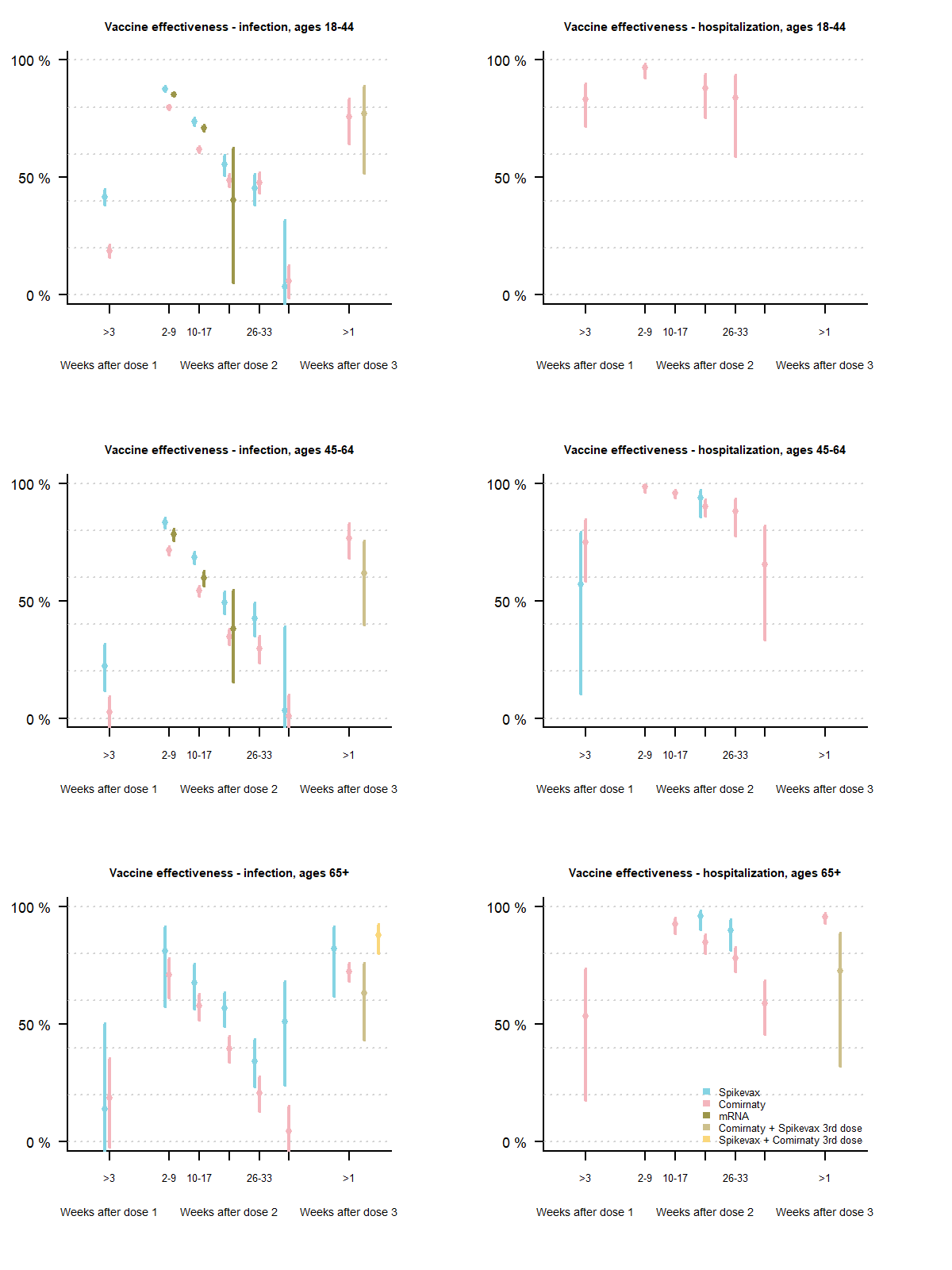

Figure S4. **Vaccine effectiveness against infection (left panels) and hospitalisations (right panels) for different vaccine products and combinations of booster doses when splitting the cohort by age.** Top panels show vaccine effectiveness estimates for ages 18-44 years, middle panels 45-64 years and lowermost panels for individuals 65 years and older. The qualitatively same pattern appears as in the analyses shown in figure 3 in the main manuscript.

Table S2. **Vaccine effectiveness against infection for all adults (18+) in Norway.** Unadjusted are given for the full cohort, adjusted (aVE) use all covariates as stratification variables in a Cox regression. Subcohort 1 excludes unvaccinated individuals who did not get a SARS-CoV-2 PCR test from December 2020 – January 2022. Subcohort 2 excludes all with comorbidities yielding a high or moderate risk of severe illness.

| **Vaccine status** | **Unadjusted VE**  **full cohort** | **aVE - full cohort** | **aVE - subcohort 1** | **Infections**  **full cohort & subcohort 1** | **Personyears**  **full cohort**  **(subcohort 1)** | **aVE - subcohort 2** | **Infections**  **subcohort 2** | **Person years**  **subcohort 2** |
| --- | --- | --- | --- | --- | --- | --- | --- | --- |
| Unvaccinated |  |  |  | 19 670 | 183 997  (128 982) |  | 18 162 | 165 757 |
| 1 week before until 3 weeks after dose 1 | 39·1 % (36·3 ‒ 41·8) | 46·0 % (43·4 ‒ 48·5) | 60·0 % (58·0 ‒ 61·8) | 2 276 | 73 784 | 45·7 % (43·0 ‒ 48·3) | 2 155 | 69 623 |
| >3 weeks after dose 1 | 18·9 % (16·7 ‒ 21·0) | 21·2 % (19·0 ‒ 23·3) | 43·3 % (41·8 ‒ 44·9) | 9 191 | 163 708 | 20·8 % (18·5 ‒ 23·1) | 8 613 | 151 116 |
| <1 week after dose 2 | 53·7 % (51·3 ‒ 56·1) | 56·0 % (53·6 ‒ 58·2) | 68·4 % (66·7 ‒ 70·0) | 1 665 | 40 638 | 55·8 % (53·4 ‒ 58·2) | 1 570 | 37 257 |
| Dose 2: 2-9 weeks | 82·8 % (82·2 ‒ 83·3) | 81·3 % (80·7 ‒ 81·9) | 87·0 % (86·6 ‒ 87·4) | 5 528 | 377 421 | 81·6 % (81·0 ‒ 82·2) | 5 003 | 327 953 |
| Dose 2: 10-17 weeks | 72·2 % (71·6 ‒ 72·8) | 63·8 % (62·9 ‒ 64·6) | 75·4 % (74·8 ‒ 76·0) | 17 189 | 389 106 | 63·9 % (63·0 ‒ 64·8) | 15 094 | 293 973 |
| Dose 2: 18-25 weeks | 66·5 % (65·7 ‒ 67·3) | 44·3 % (42·7 ‒ 45·9) | 63·3 % (62·2 ‒ 64·3) | 9 985 | 218 114 | 43·2 % (41·3 ‒ 45·0) | 6 512 | 129 799 |
| Dose 2: 26-33 weeks | 64·9 % (63·8 ‒ 65·9) | 35·1 % (32·8 ‒ 37·4) | 57·1 % (55·5 ‒ 58·6) | 5 233 | 80 587 | 35·4 % (32·7 ‒ 38·1) | 3 396 | 45 235 |
| Dose 2: >33 weeks | 49·3 % (47·0 ‒ 51·5) | 8·6 % (4·0 ‒ 13·1) | 39·2 % (36·0 ‒ 42·2) | 2 174 | 23 851 | 6·7 % (1·2 ‒ 11·8) | 1 478 | 13 724 |
| Previous infection | 92·0 % (91·0 ‒ 92·9) | 93·1 % (92·3 ‒ 93·9) | 95·2 % (94·7 ‒ 95·8) | 289 | 26 078 | 93·0 % (92·0 ‒ 93·8) | 254 | 21 840 |
| Dose 3: First week | 77·0 % (75·1 ‒ 78·7) | 45·4 % (40·6 ‒ 49·7) | 64·5 % (61·4 ‒ 67·4) | 669 | 9 540 | 42·0 % (35·5 ‒ 47·9) | 392 | 4 778 |
| Dose 3: > 1 week | 90·5 % (89·6 ‒ 91·3) | 75·6 % (73·1 ‒ 77·8) | 84·2 % (82·6 ‒ 85·7) | 502 | 16 872 | 79·1 % (75·5 ‒ 82·2) | 173 | 6 803 |

Table S3. **Vaccine effectiveness against hospitalisation for all adults (18+) in Norway.** Unadjusted are given for the full cohort, adjusted (aVE) use all covariates as stratification variables in a Cox regression. Subcohort 1 excludes unvaccinated individuals who did not get a PCR (?) test from December 2020 – January 2022. Subcohort 2 excludes all with comorbidities yielding a high or moderate risk of severe illness.

| **Vaccine status** | **Unadjusted VE**  **full cohort** | **aVE - full cohort** | **aVE - subcohort 1** | **Hospital**  **full cohort & subcohort 1** | **Person-years**  **full cohort**  **(subcohort 1)** | **aVE**  **subcohort 2** | **Hospital**  **subcohort 2** | **Person-years**  **subcohort 2** |
| --- | --- | --- | --- | --- | --- | --- | --- | --- |
| Unvaccinated |  |  |  | 652 | 186 688  (131 673) |  | 481 | 168 |
| 1 week before until 3 weeks after dose 1 | 76·5% (65·1 ‒ 84·2) | 62·3% (43·2 ‒ 74·9) | 74·3% (61·6 ‒ 82·8) | 27 | 73 892 | 66·0% (45·9 ‒ 78·7) | 21 | 69 725 |
| >3 weeks after dose 1 | 83·9% (79·0 ‒ 87·7) | 74·6% (66·6 ‒ 80·7) | 84·2% (79·2 ‒ 87·9) | 63 | 166 291 | 79·0% (70·7 ‒ 85·0) | 43 | 153 552 |
| <1 week after dose 2 |  |  |  | 4 | 40 668 |  | 3 | 37 285 |
| Dose 2: 2-9 weeks | 99·0% (98·3 ‒ 99·4) | 98·6% (97·5 ‒ 99·2) | 99·2% (98·6 ‒ 99·6) | 13 | 378 084 | 99·1% (98·1 ‒ 99·6) | 7 | 328 570 |
| Dose 2: 10-17 weeks | 96·1% (95·0 ‒ 96·9) | 96·1% (95·0 ‒ 97·0) | 97·8% (97·2 ‒ 98·3) | 71 | 390 477 | 97·8% (96·7 ‒ 98·6) | 25 | 295 163 |
| Dose 2: 18-25 weeks | 78·8% (75·1 ‒ 81·9) | 89·0% (86·7 ‒ 90·8) | 93·2% (91·8 ‒ 94·4) | 207 | 219 068 | 92·6% (89·7 ‒ 94·7) | 49 | 130 431 |
| Dose 2: 26-33 weeks | 55·2% (47·2 ‒ 62·0) | 83·9% (80·4 ‒ 86·8) | 90·2% (88·0 ‒ 92·0) | 189 | 81 074 | 90·5% (86·0 ‒ 93·5) | 37 | 45 559 |
| Dose 2: >33 weeks | -11·3% (-34·4 ‒ 7·8) | 66·6% (57·9 ‒ 73·6) | 80·2% (75·0 ‒ 84·4) | 143 | 24 095 | 73·3% (59·6 ‒ 82·3) | 36 | 13 892 |
| Previous infection |  |  |  | 4 | 24 984 |  | 2 | 21 287 |
| Dose 3: First week | 61·1% (41·6 ‒ 74·1) | 90·3% (85·2 ‒ 93·7) | 94·3% (91·3 ‒ 96·3) | 25 | 9 548 | 94·3% (85·6 ‒ 97·7) | 5 | 4 782 |
| Dose 3: > 1 week | 73·3% (61·3 ‒ 81·5) | 95·0% (92·6 ‒ 96·6) | 96·8% (95·2 ‒ 97·8) | 31 | 16 905 |  | 2 | 6 815 |

Table S4. **Vaccine effectiveness against admission to intensive care unit (ICU) for all adults (18+) in Norway.** Unadjusted are given for the full cohort, adjusted (aVE) use all covariates as stratification variables in a Cox regression. Subcohort 1 excludes unvaccinated individuals who did not get a PCR (?) test from December 2020 – January 2022. Subcohort 2 excludes all with comorbidities yielding a high or moderate risk of severe illness.

| **Vaccine status** | **Unadjusted VE**  **full cohort** | **aVE - full cohort** | **aVE - subcohort 1** | **ICU admission**  **full cohort & subcohort 1** | **Person-years**  **full cohort**  **(subcohort 1)** | **aVE**  **subcohort 2** | **ICU admissions**  **subcohort 2** | **Person-years**  **subcohort 2** |
| --- | --- | --- | --- | --- | --- | --- | --- | --- |
| Unvaccinated |  |  |  | 167 | 186 757  (131 742) |  | 101 | 168 333 |
| 1 week before until 3 weeks after dose 1 |  |  |  | 3 | 73 894 |  | 2 | 69 727 |
| >3 weeks after dose 1 | 94·3% (86·1 ‒ 97·7) | 90·1% (75·5 ‒ 96·0) | 94·7% (86·7 ‒ 97·9) | 5 | 166 309 |  | 4 | 153 567 |
| <1 week after dose 2 |  |  |  | 0 | 40 668 |  | 0 | 37 286 |
| Dose 2: 2-9 weeks |  |  |  | 1 | 378 086 |  | 1 | 328 572 |
| Dose 2: 10-17 weeks | 96·2% (93·9 ‒ 97·7) | 96·9% (94·8 ‒ 98·1) | 98·3% (97·1 ‒ 99·0) | 18 | 390 481 | 98·1% (95·5 ‒ 99·2) | 6 | 295 164 |
| Dose 2: 18-25 weeks | 84·3% (77·8 ‒ 88·9) | 94·1% (91·3 ‒ 96·0) | 96·1% (94·2 ‒ 97·4) | 41 | 219 082 | 97·3% (93·0 ‒ 99·0) | 5 | 130 435 |
| Dose 2: 26-33 weeks | 72·4% (59·3 ‒ 81·3) | 92·2% (87·8 ‒ 95·0) | 94·9% (91·9 ‒ 96·8) | 31 | 81 090 |  | 4 | 45 562 |
| Dose 2: >33 weeks | 66·1% (38·5 ‒ 81·3) | 86·6% (73·9 ‒ 93·2) | 91·7% (83·6 ‒ 95·8) | 12 | 24 110 |  | 4 | 13 896 |
| Previous infection |  |  |  | 1 | 25 934 |  | 1 | 21 794 |
| Dose 3: First week |  |  |  | 4 | 9 548 |  | 1 | 4 782 |
| Dose 3: > 1 week | 77·2% (50·5 ‒ 89·5) | 95·2% (89·0 ‒ 97·9) |  | 7 | 16 906 |  | 0 | 6 815 |

Table S5. **Vaccine effectiveness against mortality for all adults (18+) in Norway.** Unadjusted are given for the full cohort, adjusted (aVE) use all covariates as stratification variables in a Cox regression. Subcohort 1 excludes unvaccinated individuals who did not get a PCR (?) test from December 2020 – January 2022. Subcohort 2 excludes all with comorbidities yielding a high or moderate risk of severe illness.

| **Vaccine status** | **Unadjusted VE**  **full cohort** | **aVE - full cohort** | **aVE - subcohort 1** | **Deaths**  **full cohort & subcohort 1** | **Person-years**  **full cohort**  **(subcohort 1)** | **aVE - subcohort 2** | **Deaths**  **subcohort 2** | **Person-years**  **subcohort 2** |
| --- | --- | --- | --- | --- | --- | --- | --- | --- |
| Unvaccinated |  |  |  | 81 | 186 777  (131 762) |  | 32 | 168 345 |
| 1 week before until 3 weeks after dose 1 |  |  |  | 1 | 73 894 |  | 1 | 69 727 |
| >3 weeks after dose 1 | 77·2% (55·1 ‒ 88·4) | 45·2% (-6·4 ‒ 71·8) | 70·5% (42·9 ‒ 84·8) | 10 | 166 311 |  | 1 | 153 568 |
| <1 week after dose 2 |  |  |  | 1 | 40 668 |  | 1 | 37 286 |
| Dose 2: 2-9 weeks |  |  |  | 3 | 378 086 |  | 0 | 328 572 |
| Dose 2: 10-17 weeks | 96·6% (93·0 ‒ 98·4) | 93·4% (85·5 ‒ 97·0) | 96·9% (93·2 ‒ 98·6) | 8 | 390 481 |  | 3 | 295 165 |
| Dose 2: 18-25 weeks | 61·2% (44·2 ‒ 73·1) | 84·9% (77·1 ‒ 90·0) | 91·9% (87·7 ‒ 94·6) | 48 | 219 085 | 94·5 % (85·4 ‒ 97·9) | 6 | 130 435 |
| Dose 2: 26-33 weeks | -17·3% (-63·3 ‒ 15·8) | 82·1% (74·2 ‒ 87·6) | 89·9% (85·4 ‒ 93·0) | 67 | 81 092 | 80·1 % (57·8 ‒ 90·6) | 13 | 45 563 |
| Dose 2: >33 weeks | -373·5% (-559·1 ‒ -240·1) | 68·7% (55·5 ‒ 77·9) | 82·1% (74·5 ‒ 87·4) | 80 | 24 112 | 73·0 % (42·7 ‒ 87·2) | 14 | 13 896 |
| Previous infection |  |  |  | 3 | 26 116 |  | 0 | 21 874 |
| Dose 3: First week | -49·4% (-168·7 ‒ 16·9) | 88·6% (79·4 ‒ 93·7) | 93·6 % (88·5 ‒ 96·5) | 14 | 9 548 | 62·3 % (5·9 ‒ 84·9) | 7 | 4 782 |
| Dose 3: > 1 week | 10·9% (-57·9 ‒ 49·7) | 95·8% (92·6 ‒ 97·7) | 97·6 % (95·7 ‒ 98·7) | 15 | 16 907 |  | 0 | 6 816 |

Table S6. **Adjusted vaccine effectiveness (aVE) against SARS-CoV-2 infection by age groups (18-44, 45-64 and ≥65 years) among individuals in Norway, 15 July 2021 – 30 November 2021**. Result split for complete study population (full cohort) and excluding unvaccinated without any SARS-CoV-2 PCR test recorded (subcohort 1). Adjusted using strata for risk groups, county of residence, country of birth, living conditions, sex and age.

| **Vaccine status** | **aVE - full cohort** | **aVE - subcohort 1** | **Infections** | **Personyears**  **full cohort (subcohort 1)** |
| --- | --- | --- | --- | --- |
| **18-44 year old** |  |  |  |  |
| Unvaccinated | Ref | Ref | 15 085 | 122 546 (93 827) |
| 1 week before until 3 weeks after dose 1 | 48·9 % (46·3 ‒ 51·4) | 60·4 % (58·4 ‒ 62·4) | 2 025 | 62 454 |
| >3 weeks after dose 1 | 24·4 % (22·0 ‒ 26·7) | 42·8 % (41·0 ‒ 44·5) | 7 464 | 98 454 |
| <1 week after dose 2 | 58·0 % (55·5 ‒ 60·5) | 68·3 % (66·4 ‒ 70·1) | 1 313 | 24 710 |
| Dose 2: 2-9 weeks | 83·2 % (82·6 ‒ 83·8) | 87·6 % (87·1 ‒ 88·1) | 3 629 | 202 928 |
| Dose 2: 10-17 weeks | 66·4 % (65·4 ‒ 67·4) | 75·5 % (74·8 ‒ 76·2) | 9 623 | 133 290 |
| Dose 2: 18-25 weeks | 46·6 % (44·3 ‒ 48·9) | 60·8 % (59·1 ‒ 62·5) | 2 878 | 32 526 |
| Dose 2: 26-33 weeks | 40·8 % (37·3 ‒ 44·0) | 56·5 % (54·0 ‒ 58·9) | 1 434 | 11 001 |
| Dose 2: >33 weeks | 5·2 % (-1·9 ‒ 11·8) | 30·7 % (25·5 ‒ 35·5) | 828 | 4 309 |
| Dose 3: First week | 36·1 % (22·1 ‒ 47·5) | 53·4 % (43·2 ‒ 61·8) | 101 | 448 |
| Dose 3: > 1 week | 76·5 % (67·9 ‒ 82·8) | 82·8 % (76·5 ‒ 87·4) | 40 | 655 |
| Previous infection | 92·6 % (91·5 ‒ 93·5) | 94·5 % (93·8 ‒ 95·2) | 223 | 15 585 |
| **45-64 year old** |  |  |  |  |
| Unvaccinated | Ref | Ref | 3 930 | 44 019 (27 286) |
| 1 week before until 3 weeks after dose 1 | 25·5 % (14·6 ‒ 35·0) | 53·2 % (46·4 ‒ 59·1) | 234 | 10 465 |
| >3 weeks after dose 1 | 8·4 % (2·2 ‒ 14·2) | 42·7 % (38·9 ‒ 46·3) | 1 632 | 60 959 |
| <1 week after dose 2 | 49·2 % (42·9 ‒ 54·9) | 68·3 % (64·4 ‒ 71·8) | 337 | 15 280 |
| Dose 2: 2-9 weeks | 75·6 % (74·1 ‒ 77·0) | 85·0 % (84·1 ‒ 85·9) | 1 827 | 147 044 |
| Dose 2: 10-17 weeks | 57·4 % (55·5 ‒ 59·2) | 74·3 % (73·2 ‒ 75·4) | 6 897 | 151 677 |
| Dose 2: 18-25 weeks | 36·4 % (33·3 ‒ 39·4) | 61·8 % (59·9 ‒ 63·7) | 4 444 | 61 704 |
| Dose 2: 26-33 weeks | 32·7 % (28·2 ‒ 37·0) | 59·5 % (56·8 ‒ 62·1) | 1 454 | 13 546 |
| Dose 2: >33 weeks | 0·5 % (-9·4 ‒ 9·5) | 40·1 % (34·1 ‒ 45·5) | 514 | 4 175 |
| Dose 3: First week | 33·1 % (17·8 ‒ 45·6) | 59·8 % (50·6 ‒ 67·3) | 95 | 769 |
| Dose 3: > 1 week | 72·1 % (64·7 ‒ 77·9) | 83·2 % (78·8 ‒ 86·7) | 74 | 1 720 |
| Previous infection | 95·1 % (93·6 ‒ 96·3) | 97·0 % (96·1 ‒ 97·7) | 52 | 8 084 |
| **≥ 65 years old** |  |  |  |  |
| Unvaccinated | Ref | Ref | 655 | 17 432 (7 869) |
| 1 week before until 3 weeks after dose 1 | 31·9 % (-10·5 ‒ 58·0) | 70·2 % (51·7 ‒ 81·6) | 17 | 865 |
| >3 weeks after dose 1 | 19·3 % (-0·2 ‒ 35·0) | 64·7 % (56·2 ‒ 71·6) | 95 | 4 296 |
| <1 week after dose 2 | 2·7 % (-62·8 ‒ 41·8) | 57·2 % (28·4 ‒ 74·4) | 15 | 648 |
| Dose 2: 2-9 weeks | 74·9 % (67·2 ‒ 80·7) | 88·9 % (85·5 ‒ 91·5) | 72 | 27 448 |
| Dose 2: 10-17 weeks | 59·1 % (53·5 ‒ 64·0) | 82·0 % (79·5 ‒ 84·1) | 669 | 104 140 |
| Dose 2: 18-25 weeks | 41·5 % (35·9 ‒ 46·7) | 74·7 % (72·2 ‒ 76·9) | 2 663 | 123 884 |
| Dose 2: 26-33 weeks | 22·4 % (14·8 ‒ 29·3) | 66·3 % (63·0 ‒ 69·3) | 2 345 | 56 040 |
| Dose 2: >33 weeks | 8·4 % (-2·8 ‒ 18·5) | 59·9 % (55·0 ‒ 64·3) | 832 | 15 367 |
| Dose 3: First week | 43·3 % (35·7 ‒ 50·0) | 75·4 % (72·1 ‒ 78·3) | 473 | 8 324 |
| Dose 3: > 1 week | 73·7 % (69·8 ‒ 77·0) | 88·5 % (86·9 ‒ 90·0) | 388 | 14 497 |
| Previous infection | 89·0 % (81·3 ‒ 93·5) | 95·2 % (91·8 ‒ 97·2) | 14 | 2 408 |

Table S7. **Adjusted vaccine effectiveness (aVE) against hospitalisation by age groups (18-44, 45-64 and ≥65 years) among individuals in Norway, 15 July 2021 – 30 November 2021.** Result split for complete study population (full cohort) and excluding unvaccinated without any SARS-CoV-2 PCR test recorded (subcohort 1). Adjusted using strata for risk groups, county of residence, country of birth, living conditions, sex and age.

| **Vaccine status** | **aVE - full cohort** | **aVE - subcohort 1** | **Hospitalisations** | **Personyears**  **full cohort (subcohort 1)** |
| --- | --- | --- | --- | --- |
| **18–44-year-old** |  |  |  |  |
| Unvaccinated | Ref | Ref | 253 | 124 757 (96 038) |
| 1 week before until 3 weeks after dose 1 | 75·1 % (55·0 ‒ 86·2) | 80·6 % (65·0 ‒ 89·2) | 13 | 62 552 |
| >3 weeks after dose 1 | 82·9 % (73·0 ‒ 89·2) | 87·0 % (79·4 ‒ 91·7) | 22 | 100 596 |
| <1 week after dose 2 | n.a. | n.a. | 1 | 24 735 |
| Dose 2: 2-9 weeks | 98·4 % (96·4 ‒ 99·3) | 98·8 % (97·3 ‒ 99·5) | 6 | 203 405 |
| Dose 2: 10-17 weeks | 98·3 % (96·4 ‒ 99·2) | 98·8 % (97·4 ‒ 99·4) | 7 | 134 065 |
| Dose 2: 18-25 weeks | 89·8 % (81·4 ‒ 94·5) | 92·3 % (86·0 ‒ 95·8) | 13 | 32 833 |
| Dose 2: 26-33 weeks | 86·7 % (70·3 ‒ 94·1) | 89·9 % (77·5 ‒ 95·5) | 7 | 11 146 |
| Dose 2: >33 weeks | n.a. | n.a. | 3 | 4 398 |
| Dose 3: First week | n.a. | n.a. | 0 | 449 |
| Dose 3: > 1 week | n.a. | n.a. | 1 | 658 |
| Previous infection | n.a. | n.a. | 1 | 15 390 |
| **45–64-year-old** |  |  |  |  |
| Unvaccinated | Ref | Ref | 239 | 44 450 (27 717) |
| 1 week before until 3 weeks after dose 1 | 38·4 % (-14·5 ‒ 66·9) | 60·5 % (26·7 ‒ 78·7) | 11 | 10 474 |
| >3 weeks after dose 1 | 71·4 % (56·2 ‒ 81·4) | 81·9 % (72·3 ‒ 88·2) | 27 | 61 387 |
| <1 week after dose 2 | n.a. | n.a. | 1 | 15 285 |
| Dose 2: 2-9 weeks | 99·1 % (97·7 ‒ 99·6) | 99·4 % (98·6 ‒ 99·8) | 5 | 147 227 |
| Dose 2: 10-17 weeks | 96·4 % (94·7 ‒ 97·5) | 97·9 % (97·0 ‒ 98·6) | 35 | 152 227 |
| Dose 2: 18-25 weeks | 90·9 % (87·4 ‒ 93·5) | 94·8 % (92·7 ‒ 96·3) | 57 | 62 141 |
| Dose 2: 26-33 weeks | 91·5 % (84·7 ‒ 95·2) | 95·0 % (91·1 ‒ 97·2) | 14 | 13 687 |
| Dose 2: >33 weeks | 65·3 % (33·3 ‒ 82·0) | 80·5 % (62·3 ‒ 89·9) | 10 | 4 232 |
| Dose 3: First week | n.a. | n.a. | 1 | 770 |
| Dose 3: > 1 week | 85·4 % (65·3 ‒ 93·9) | 91·5 % (79·8 ‒ 96·4) | 6 | 1 725 |
| Previous infection | n.a. | n.a. | 2 | 7 589 |
| **≥ 65 years old** |  |  |  |  |
| Unvaccinated | Ref | Ref | 160 | 17 481 (7 917) |
| 1 week before until 3 weeks after dose 1 | n.a. | n.a. | 3 | 866 |
| >3 weeks after dose 1 | 56·5 % (24·6 ‒ 74·9) | 79·2 % (64·0 ‒ 88·0) | 14 | 4 307 |
| <1 week after dose 2 | n.a. | n.a. | 2 | 649 |
| Dose 2: 2-9 weeks | n.a. | n.a. | 2 | 27 452 |
| Dose 2: 10-17 weeks | 93·6 % (89·9 ‒ 96·0) | 96·9 % (95·2 ‒ 98·1) | 29 | 104 185 |
| Dose 2: 18-25 weeks | 86·2 % (82·2 ‒ 89·4) | 93·5 % (91·5 ‒ 94·9) | 137 | 124 093 |
| Dose 2: 26-33 weeks | 79·9 % (74·5 ‒ 84·1) | 90·4 % (87·9 ‒ 92·4) | 168 | 56 240 |
| Dose 2: >33 weeks | 61·9 % (50·1 ‒ 70·9) | 81·7 % (76·1 ‒ 86·0) | 130 | 15 465 |
| Dose 3: First week | 88·8 % (82·5 ‒ 92·8) | 94·7 % (91·7 ‒ 96·6) | 24 | 8 329 |
| Dose 3: > 1 week | 95·3 % (92·6 ‒ 97·0) | 97·8 % (96·5 ‒ 98·6) | 24 | 14 522 |
| Previous infection | n.a. | n.a. | 1 | 2 005 |

Table S8. **Adjusted vaccine effectiveness (aVE) against SARS-CoV-2 infection and hospitalisation by vaccine product type among individuals in Norway, 15 July 2021 – 30 November 2021.** Adjusted using strata for risk groups, county of residence, country of birth, living conditions, sex and age.

|  |  | **SARS-CoV-2 infections** | | | **Hospitalisations** | | |
| --- | --- | --- | --- | --- | --- | --- | --- |
|  | **Vaccine status** | **Adjusted VE** | **Events** | **Person years** | **Adjusted VE** | **Events** | **Person years** |
| **Comirnaty** | |  |  |  |  |  |  |
|  | Unvaccinated | n.a. | 19 670 | 183 997 | n.a. | 652 | 186 688 |
|  | 1 week before until 3 weeks after dose 1 | 43·7 % (40·7 ‒ 46·6) | 1 823 | 60 651 | 58·8 % (34·5 ‒ 74·1) | 21 | 60 735 |
|  | >3 weeks after dose 1 | 15·5 % (12·9 ‒ 18·0) | 7 532 | 133 878 | 75·3 % (66·4 ‒ 81·9) | 48 | 136 031 |
|  | <1 week after dose 2 | 52·8 % (49·4 ‒ 56·0) | 891 | 23 179 | n.a. | 3 | 23 196 |
|  | Dose 2: 2-9 weeks | 77·7 % (76·8 ‒ 78·5) | 3 352 | 231 500 | 97·5 % (95·6 ‒ 98·6) | 13 | 231 883 |
|  | Dose 2: 10-17 weeks | 60·0 % (58·9 ‒ 61·0) | 10 998 | 278 007 | 95·4 % (93·9 ‒ 96·5) | 62 | 278 910 |
|  | Dose 2: 18-25 weeks | 43·9 % (42·1 ‒ 45·7) | 7 509 | 176 006 | 87·5 % (84·8 ‒ 89·7) | 189 | 176 680 |
|  | Dose 2: 26-33 weeks | 34·3 % (31·4 ‒ 37·1) | 3 429 | 67 758 | 81·8 % (77·7 ‒ 85·2) | 172 | 68 071 |
|  | Dose 2: >33 weeks | 8·2 % (3·4 ‒ 12·8) | 2 100 | 23 227 | 63·9 % (54·3 ‒ 71·5) | 141 | 23 461 |
| **Spikevax** | |  |  |  |  |  |  |
|  | 1 week before until 3 weeks after dose 1 | 50·0 % (45·0 ‒ 54·5) | 452 | 13 119 | 70·8 % (34·0 ‒ 87·0) | 6 | 13 142 |
|  | >3 weeks after dose 1 | 38·1 % (34·7 ‒ 41·3) | 1 579 | 28 914 | 77·1 % (58·9 ‒ 87·3) | 12 | 29 332 |
|  | <1 week after dose 2 | 67·7 % (62·7 ‒ 72·0) | 192 | 5 302 | n.a. | 0 | 5 305 |
|  | Dose 2: 2-9 weeks | 86·6 % (85·6 ‒ 87·6) | 753 | 46 456 | n.a. | 0 | 46 540 |
|  | Dose 2: 10-17 weeks | 72·3 % (70·9 ‒ 73·5) | 2 170 | 43 097 | n.a. | 4 | 43 268 |
|  | Dose 2: 18-25 weeks | 55·2 % (52·4 ‒ 57·9) | 1 193 | 21 826 | 95·3 % (91·5 ‒ 97·4) | 12 | 21 962 |
|  | Dose 2: 26-33 weeks | 44·4 % (40·1 ‒ 48·3) | 835 | 7 588 | 91·1 % (84·9 ‒ 94·8) | 15 | 7 665 |
|  | Dose 2: >33 weeks | 28·6 % (9·6 ‒ 43·6) | 72 | 603 | n.a. | 1 | 613 |
| **mRNA combined** | |  |  |  |  |  |  |
|  | <1 week after dose 2 | 53·7 % (49·6 ‒ 57·5) | 579 | 12 098 | n.a. | 1 | 12 107 |
|  | Dose 2: 2-9 weeks | 84·1 % (83·2 ‒ 85·0) | 1 386 | 95 094 | n.a. | 0 | 95 288 |
|  | Dose 2: 10-17 weeks | 68·5 % (67·2 ‒ 69·7) | 3 495 | 47 688 | n.a. | 4 | 47 950 |
|  | Dose 2: 18-25 weeks | 40·7 % (23·9 ‒ 53·8) | 64 | 451 | n.a. | 0 | 454 |
|  | Dose 2: 26-33 weeks | n.a. | 3 | 36 | n.a. | 0 | 36 |
|  | Dose 2: >33 weeks | n.a. | 1 | 9 | n.a. | 0 | 10 |

Table S9. **Adjusted vaccine effectiveness (aVE) against SARS-CoV-2 infection and hospitalisation after receiving a booster dose by vaccine product type among individuals in Norway, 15 July 2021 – 30 November 2021.** Adjusted using strata for risk groups, county of residence, country of birth, living conditions, sex, and age.

|  |  |  | **SARS-CoV-2 infections** | | | **Hospitalisations** | | |
| --- | --- | --- | --- | --- | --- | --- | --- | --- |
| **Primary** | **Booster** |  | **Adjusted VE** | **Events** | **Person years** | **Adjusted VE** | **Events** | **Person years** |
| **Comirnaty** | Comirnaty | Dose 3: 1st week | 45·3 % (39·8 ‒ 50·2) | 515 | 7 934 | 89·1 % (83·0 ‒ 93·0) | 23 | 7 940 |
|  |  | Dose 3: >1 week | 75·3 % (72·5 ‒ 77·8) | 408 | 13 994 | 95·6 % (93·1 ‒ 97·2) | 22 | 14 021 |
|  | Spikevax | Dose 3: 1st week | 39·9 % (22·0 ‒ 53·7) | 58 | 601 | n.a. | 2 | 602 |
|  |  | Dose 3: >1 week | 68·2 % (57·6 ‒ 76·1) | 48 | 1 137 | 73·5 % (45·7 ‒ 87·1) | 8 | 1 140 |
| **Spikevax** | Spikevax | Dose 3: 1st week | 56·1 % (33·0 ‒ 71·2) | 22 | 265 | n.a. | 0 | 265 |
|  |  | Dose 3: >1 week | 84·9 % (71·8 ‒ 91·9) | 10 | 354 | n.a. | 1 | 355 |
|  | Comirnaty | Dose 3: 1st week | 67·6 % (48·4 ‒ 79·7) | 18 | 423 | n.a. | 0 | 423 |
|  |  | Dose 3: >1 week | 87·1 % (80·1 ‒ 91·6) | 21 | 1 135 | n.a. | 0 | 1 136 |
